# Morphology of proximal and distal human semitendinosus compartments and the effects of distal tendon harvesting for anterior cruciate ligament reconstruction

**DOI:** 10.1101/2022.07.22.22277919

**Authors:** Adam Kositsky, Huub Maas, Rod S. Barrett, Ben Kennedy, Lauri Stenroth, Rami K. Korhonen, Chris J. Vertullo, Laura E. Diamond, David J. Saxby

## Abstract

The human semitendinosus muscle is characterized by a tendinous inscription separating proximal (ST_prox_) and distal (ST_dist_) neuromuscular compartments. As each compartment is innervated by separate nerve branches, potential exists for the compartments to operate and be controlled independently. However, the morphology and function of each compartment have not been thoroughly examined in a human adult population. Further, the distal semitendinosus tendon is typically harvested for use in anterior cruciate ligament reconstruction (ACLR) surgery, which induces long-term morphological changes to the semitendinosus muscle-tendon unit. It remains unknown if muscle morphological alterations following ACLR are uniform between ST_prox_ and ST_dist_. Here, we performed magnetic resonance imaging on ten individuals who had undergone ACLR involving an ipsilateral distal semitendinosus tendon graft 14 ± 6 months prior, extracting morphological parameters of the whole ST muscle and each individual muscle compartment from both the (non-injured) contralateral and surgical legs. In the contralateral non-surgical leg, volume and length of ST_prox_ were lower than ST_dist_. No between-compartment differences in volume or length were found for ACLR legs, likely due to greater shortening of ST_dist_ compared to ST_prox_ after ACLR. The maximal anatomical cross-sectional area of both compartments was substantially smaller on the ACLR leg, but did not differ between ST_prox_ and ST_dist_ on either leg. The absolute and relative differences in ST_prox_ morphology on the ACLR leg were strongly correlated with the corresponding between-leg differences in ST_dist_ morphological parameters. Specifically, greater morphological differences in one compartment were highly correlated with large differences in the other compartment, and vice versa for smaller differences. These relationships indicate that despite the heterogeneity in compartment length and volume, compartment atrophy is not independent or random. Further, the tendinous inscription endpoints were generally positioned at the same proximodistal level as the compartment maximal anatomical cross-sectional areas, providing a wide area over which the tendinous inscription could mechanically interact with compartments. Overall, results suggest the two human semitendinosus compartments are not mechanically independent.

## INTRODUCTION

The musculus semitendinosus (ST) is a posterior thigh muscle and one of four human hamstring muscles. The ST muscle’s culinary importance has made it an oft-studied muscle in food science (Satorius & Child, 1938; Machlik & Draudt, 1963; Purslow, 1985; Wang *et al*., 2022), but ST has also been widely used as a model to study muscle structure, composition, function, and mechanics across several non-human species (Délèze, 1961; Sivachelvan & Davies, 1981; Street, 1983; Roy *et al*., 1984; Edgerton *et al*., 1987; Gans *et al*., 1989; Kawakami & Lieber, 2000; Shimada *et al*., 2004). The ST of many, but not all (Appleton, 1928), species is characterized by the presence of a tendinous inscription (TI) that separates the ST into proximal (ST_prox_) and distal (ST_dist_) neuromuscular compartments, each containing separate nerve innervations (Hopwood & Butterfield, 1976; Roy *et al*., 1984; Edgerton *et al*., 1987; Gans *et al*., 1989; Paul, 2001; Woodley & Mercer, 2005). Despite potential for their asynchronous activations, ST_prox_ and ST_dist_ in cats are mechanically linked and generally functioning in-series, i.e., as a single muscle (Bodine *et al*., 1982; Edgerton *et al*., 1987; English & Weeks, 1987; Hutchison *et al*., 1989; Chanaud *et al*., 1991). A recent study found no differences in passive mechanical properties between ST compartments in humans, suggesting human ST_prox_ and ST_dist_ also function mechanically in-series (Kositsky *et al*., 2022). However, the morphology of ST compartments has not been well documented in humans *in vivo*. As a muscle’s structure defines its function (Bamman *et al*., 2000; Fukunaga *et al*., 2001), evaluating compartment morphology would provide further insight into the interaction between ST compartments.

Studies examining ST compartment morphology and structure in humans have generally been restricted to cadaveric investigations, which have reported conflicting results regarding differences, or lack thereof, in fascicle or fiber length (Markee *et al*., 1955; Barrett, 1962; Wickiewicz *et al*., 1983; Woodley & Mercer, 2005; Kellis *et al*., 2012; Haberfehlner *et al*., 2016*b*) and physiological cross-sectional area (Woodley & Mercer, 2005; Haberfehlner *et al*., 2016*b*) between human ST_prox_ and ST_dist_. Regarding volume, Haberfehlner *et al*. (2016*b*) found no difference between ST compartments. Although allowing for more direct measurements, cadaveric studies are typically not well standardized across investigations and generally consist of older specimens that have a high chance of being affected by neuromuscular disease or impairment. Hence, the translation of information obtained from cadavers to living healthy adults may be limited. *In vivo* studies (Haberfehlner *et al*., 2016*a*; Hanssen *et al*., 2021) have used three-dimensional freehand ultrasound to assess ST compartmental fascicle length, volume, and echo intensity (related to muscle quality) in typically developing children and those with spasticity. However, ST compartment morphology of healthy human adults has not been studied in detail. Further, quantitative examinations of the TI in human cadavers have been limited to detailing the dimensions, angle, and location along whole ST muscle length (Markee *et al*., 1955; Lee *et al*., 1988; Garrett *et al*., 1989; Woodley & Mercer, 2005; Kellis *et al*., 2012; van der Made *et al*., 2015), while *in vivo* ultrasound-based studies additionally quantified TI location only with respect to the ischial tuberosity (Kellis *et al*., 2012; Kellis & Balidou, 2014). As forces may be transferred across ST compartments (Bodine *et al*., 1982; Edgerton *et al*., 1987), the location of the TI within and between compartments may play an important role in any potential force transmission function. However, to our best knowledge, the positioning of the TI in relation to ST compartment morphology has yet to be documented.

In orthopaedics, the distal ST tendon is routinely harvested as autologous graft tissue, particularly for anterior cruciate ligament reconstruction (ACLR) (Thaunat *et al*., 2019; Vertullo *et al*., 2019). Although this surgical procedure effectively sacrifices ST, the ST tendon demonstrates remarkable potential to regenerate and reattach below the knee joint line (Nakamae *et al*., 2005; Papalia *et al*., 2015), regaining some level of function. However, after ACLR, the ST muscle belly is substantially shorter, with decreased anatomical cross-sectional area (ACSA) and volume (Williams *et al*., 2004; Makihara *et al*., 2006; Snow *et al*., 2012; Nomura *et al*., 2015; Konrath *et al*., 2016; Messer *et al*., 2020; Morris *et al*., 2021). Recently, de Moulin *et al*. (2022) observed non-uniform morphological adaptations along the ST muscle belly after ACLR, but only defined muscle regions as thirds along muscle belly length and not as anatomical compartments. The shape of ST, particularly distally, has also been qualitatively (Snow *et al*., 2012) and quantitatively (du Moulin *et al*., 2022) observed to be different after tendon harvesting for ACLR. However, it remains unknown if ST_prox_ and ST_dist_ are altered heterogeneously post-ACLR.

Differences in morphological adaptations between ST muscle compartments after ACLR might be expected as the proximal tendon and muscle portion, which remain mechanically connected to surrounding tissues, could still contribute to hip and knee joint torques (Maas & Sandercock, 2008; de Bruin *et al*., 2011). If so, ST_prox_ would experience greater loading compared to distal areas, at least until the distal tendon may reattach. Lower loading in ST_dist_ could lead to a greater reduction in compartment size and length (Wisdom *et al*., 2015; Franchi *et al*., 2022) compared to ST_prox_. Potential non-uniform morphological changes between ST_prox_ and ST_dist_, as shown in musculoskeletal conditions other than ACLR (e.g., children with cerebral palsy; Haberfehlner *et al*. [2016*a*]), may disrupt the mechanical interplay between ST compartments seen in healthy legs (Kositsky *et al*., 2022). Moreover, as ST_prox_ may function primarily at the hip, and ST_dist_ predominantly at the knee (Markee *et al*., 1955), compartment specific mechanical impairment may be relevant for the common and persistent knee flexion weakness following ACLR with an ST graft, even in the presence of ST tendon regeneration (Nakamae *et al*., 2005; Makihara *et al*., 2006; Nomura *et al*., 2015; Papalia *et al*., 2015; Konrath *et al*., 2016).

In this study, we used magnetic resonance imaging (MRI) to bilaterally evaluate the morphology of ST, including ST_prox_ and ST_dist_, in adults with a unilateral ACLR involving a distal ST tendon autograft. Specifically, we aimed to assess whole muscle and compartment morphology on the contralateral (non-surgical) leg, the effects distal ST tendon harvesting has on ST gross morphology, and how compartments may atrophy with respect to each other, the whole muscle, and the TI. We also aimed to describe the positioning of the TI in relation to compartment and whole muscle morphology and if the positioning may be affected by the morphological changes induced by ACLR.

## METHODS

### Participants

Ten participants (six females; age: 27.2 ± 4.9 years; height: 171.6 ± 10.0 cm; mass: 72.6 ± 13.4 kg; 424 ± 109 days post-surgery) were recruited for the study. All participants, of which five had accompanying meniscal lesions, underwent ACLR with a quadrupled ipsilateral semitendinosus autograft (see *Surgical procedures* section). Exclusion criteria consisted of: ACLR >6 months post initial injury, concomitant harvesting of gracilis tendon for ACLR, previous major knee injuries, neurological disorders, and/or contraindications for MRI scans. Participants were requested to refrain from strenuous exercise commencing 24 hours prior to the investigation and provided written informed consent prior to any involvement in the study. The Griffith University Human Research Ethics Committee (2018/839) approved the study, which was carried out in accordance with the Declaration of Helsinki.

### Surgical procedures

A fellowship trained orthopaedic surgeon (C.J.V.) performed all ACLRs. After application of a tourniquet to the thigh, an anteromedial vertical incision was made over the pes anserinus. The sartorius fascia was then incised to visualise the ST tendon. The tendon was left secured to the distal attachment point and an open-ended tendon harvester (Linvatec, Florida, USA) was used to release the entire distal tendon length from its muscular attachment. Then, the ST tendon was removed from its distal bony attachment with a scalpel. A quadrupled ST graft was formed using a wrapping technique over two Tightrope fixation devices (Arthrex, Florida, USA), proximally and distally, and then sutured using Fibrewire (Arthrex, Florida, USA) (Vertullo *et al*., 2019). The femoral tunnel was created via a transportal drilling technique and the tibial tunnel drilled outside-in. Femoral and tibial fixation with the adjustable fixation devices were undertaken in full extension.

### Magnetic resonance imaging acquisition and data analyses

With the participant lying supine, T_1_ Dixon three-dimensional fast field echo and two-dimensional proton density magnetic resonance images were acquired with a 3T MRI unit (Ingenia, Phillips, Eindhoven, Netherlands). Scan acquisition parameters are summarized in Table 1. For T_1_ Dixon scans, a B1 field map (dual repetition time) was used to minimize signal contrast variation across the field-of-view. Coronal T_1_ Dixon images were reconstructed into 691 axial slices (1 mm slice thickness) with in-plane pixel resolution of 0.446 mm using Mimics software (Version 20.0, Materialise, Leuven, Belgium). Each compartment was separately traced in every ∼5 axial slices in the water in-phase images, with software interpolation used for slices in between. Caution was taken to include as little of the muscle border as possible, and images were manually inspected to ensure interpolation did not cause substantial errors. The ST_prox_ and ST_dist_ masks were also combined and gaps between them filled (i.e., to include the TI, as this is how ST is typically segmented) to create a whole ST mask. Compartment and muscle belly length were calculated in the proximodistal axis by multiplying slice thickness (1 mm) by the number of slices in which the respective compartment/muscle was visible (Fukunaga *et al*., 2001; Messer *et al*., 2020). The slice containing each compartment’s and the entire muscle’s largest cross-sectional value was deemed the compartment/muscle maximal ACSA (ACSA_max_) (Fukunaga *et al*., 2001; Kositsky *et al*., 2020). The location of compartment and muscle ACSA_max_ relative to the respective compartment and entire muscle belly length was also determined, with the proximal end of the muscle corresponding to 0% and the distal end to 100%. Compartment and muscle volumes were calculated by multiplying slice thickness (1 mm) by the sum of contiguous ACSAs (Fukunaga *et al*., 2001; Messer *et al*., 2020). The position of the proximal (TI_prox_) and distal (TI_dist_) endpoints of the TI was determined relative to the length of each compartment and the entire muscle belly, and the proximodistal length (in the axial imaging plane) of the TI was determined from the number of slices in which ST_prox_ and ST_dist_ overlapped. Examples of all morphological analyses are depicted in Figure 1. The distal ST tendon was considered as regenerated if a tendinous structure was visible on proton density and T_1_ Dixon scans below the knee joint.

**Table 1.** Acquisition parameters for magnetic resonance imaging scans.

|  | **T<sub>1</sub> Dixon** | **Proton density** |
| --- | --- | --- |
| **Main purpose** | Muscle/compartiment segmentation | Visualize ST tendon |
| **Acquisition plane** | Coronal (3D) | Axial (2D) |
| **Number of stations** | Two | One |
| **Number of slices** | 252 | 80-95 |
| **Slice thickness** | 1 mm | 3 mm |
| **Slice gap** | 0 mm | 0.3 mm |
| **Station overlap** | 30 mm | - |
| **Repetition time** | 7.45 ms | 2853-3804 ms * |
| **Echo time(s)** | 1.19, 2.37 ms | 25 ms |
| **Flip angle** | 10° | 90° |
| **Voxel size** | 1 x 1 x 1 mm | 0.8 x 0.88 mm |
| **Minimum FOV** | 360 x 450 x 252 mm | 200 x 380 mm |
| **Acceleration factor** | SENSE 2 | SENSE 2.5 |
| **Acquisition time** | 14 min (7 min per station) | 5-7 min * |

FOV = field of view; ST = semitendinosus; 2D = two-dimensional; 3D = three-dimensional.
\*depending on the FOV and the number of slices

**Figure 1.**
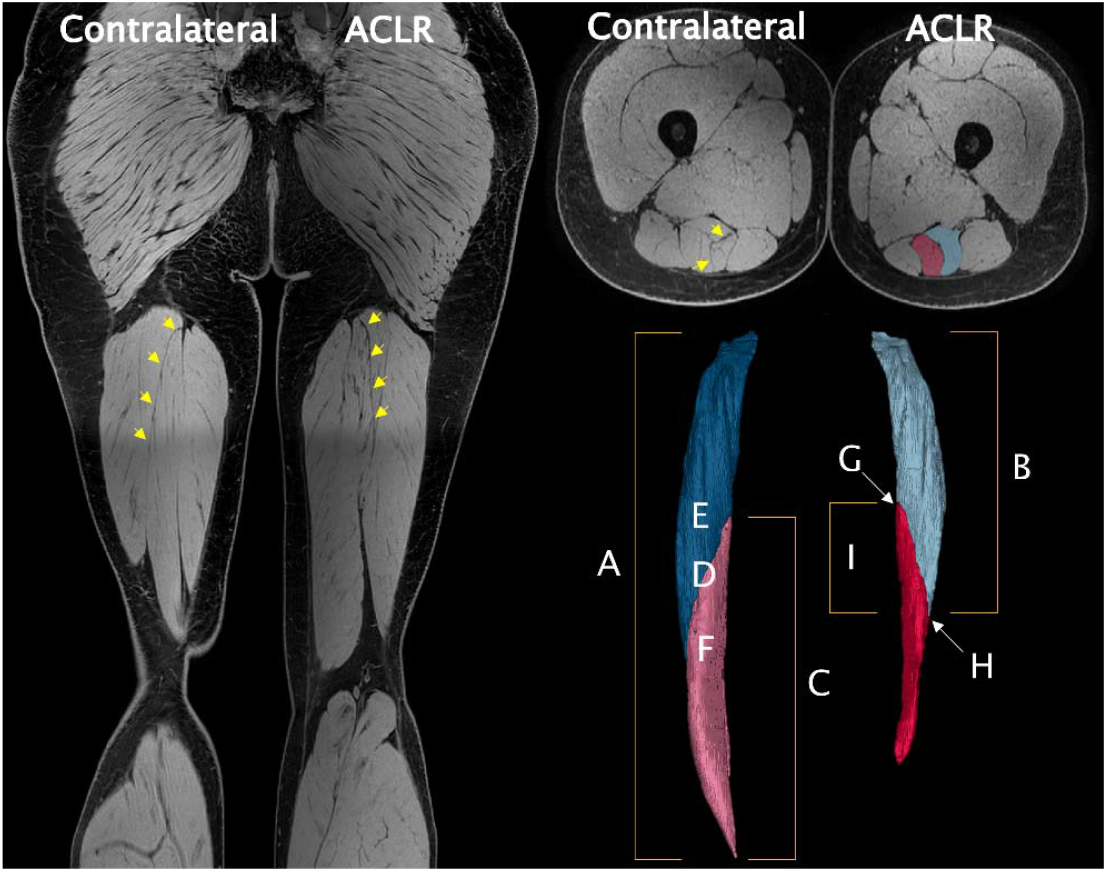
Raw coronal water in-phase magnetic resonance imaging (MRI) sequence (left). Axially reconstructed MRI image with segmentations of proximal (ST_prox_) and distal (ST_dist_) semitendinosus (ST) compartments overlayed on the anterior cruciate ligament (ACLR) leg (upper right). Example reconstruction of proximal (contralateral: dark blue; anterior cruciate ligament reconstructed: light blue) and distal (contralateral: pink; anterior cruciate ligament reconstructed: red) semitendinosus compartments (lower right). Note the full length of the ST muscle is not seen in the coronal slice, reconstructed segmentations are not scaled to the coronal image, and all images are from the same participant, who had ST tendon regeneration and 7.2 cm of muscle shortening. The tendinous inscription (TI) is indicated with yellow arrows on MRI images. *A*: ST whole muscle length. *B*: ST_prox_ compartment length. *C*: ST_dist_ compartment length. *D*: ST whole muscle ACSA_max_. *E*: ST_prox_ compartment ACSA_max_. *F*: ST_dist_ compartment ACSA_max_. *G*: proximal endpoint of TI. *H*: distal endpoint of TI. *I*: TI length.

### Statistical analyses

Paired samples t-tests were used to assess the between-leg differences in whole ST muscle morphology (volume, ACSA_max_, length), TI length, and the location of whole ST muscle ACSA_max_ relative to whole muscle and TI lengths. The effects of compartment (ST_prox_, ST_dist_) and leg (contralateral, ACLR) on volume, ACSA_max_, and length were assessed using full-factorial, two-way repeated measured ANOVAs. To confirm grouping all participants in a single cohort regardless of tendon regeneration status did not affect our results, the paired samples t-tests and two-way repeated measured ANOVAs for volume, ACSA_max_, and length were repeated with only tendon regenerated participants included. A two-way repeated measures ANOVA was performed to assess if the location of ST_prox_ ACSA_max_ relative to muscle length changed after ACLR or differed from TI_prox_ across legs. A separate ANOVA was performed comparing the locations of ST_dist_ ACSA_max_ and TI_dist_. For all repeated measures ANOVAs, Bonferroni corrections were applied when significant interactions were found. The between-leg differences for each compartment morphological parameter (e.g., volume of ST_prox_ on the ACLR leg minus volume of ST_prox_ on the contralateral leg) were determined and Pearson’s *r* correlation coefficients used to assess the relationships in between-leg differences for (i) ST_prox_ and ST_dist_ morphology, (ii) compartment and whole muscle morphology, and (iii) compartment and TI length. All statistical analyses were performed with SPSS (v27, SPSS Inc., Chicago, IL, USA). All data are presented as mean ± one standard deviation.

## RESULTS

Distal ST tendon regeneration was observed in seven of the ten participants. Whole ST muscle volume (p < 0.001), ACSA_max_ (p = 0.02), and length (p = 0.001) were all smaller on the ACLR compared to contralateral leg (Table 2). The location of whole muscle ACSA_max_ relative to whole muscle length was more distal on the ACLR leg (contralateral: 40.8 ± 3.3%; ACLR: 49.0 ± 8.4%; p = 0.025), but the location of ACSA_max_ along the TI length did not differ across legs (contralateral: 36.5 ± 12.0%; ACLR: 39.5 ± 15.8%; p = 0.623).

**Table 2.**
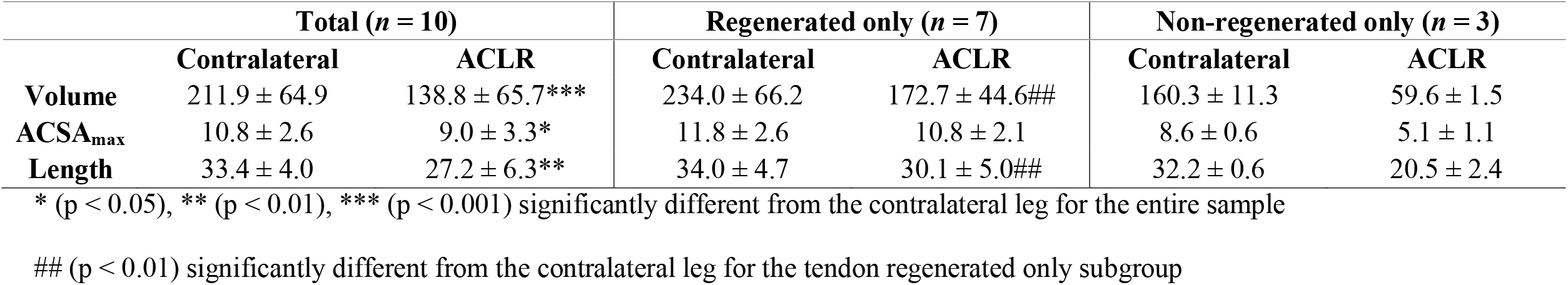
Means and standard deviations of volume, maximal anatomical cross-sectional area (ACSA_max_), and length of the whole semitendinosus muscle for contralateral and anterior cruciate ligament reconstructed (ACLR) legs. Paired samples t-tests for between-leg differences were performed for the entire sample and for the tendon regenerated only subgroup, and not for the non-regenerated only subgroup due to sample size.

|  | **Total (*n* = 10)** |  | **Regenerated only (*n* = 7)** |  | **Non-regenerated only (*n* = 3)** |  |
| --- | --- | --- | --- | --- | --- | --- |
|  | **Contralateral** | **ACLR** | **Contralateral** | **ACLR** | **Contralateral** | **ACLR** |
| **Volume** | 211.9 ± 64.9 | 138.8 ± 65.7*** | 234.0 ± 66.2 | 172.7 ± 44.6## | 160.3 ± 11.3 | 59.6 ± 1.5 |
| **ACSA<sub>max</sub>** | 10.8 ± 2.6 | 9.0 ± 3.3* | 11.8 ± 2.6 | 10.8 ± 2.1 | 8.6 ± 0.6 | 5.1 ± 1.1 |
| **Length** | 33.4 ± 4.0 | 27.2 ± 6.3** | 34.0 ± 4.7 | 30.1 ± 5.0## | 32.2 ± 0.6 | 20.5 ± 2.4 |

\* ( $p < 0.05$ ), \*\* ( $p < 0.01$ ), \*\*\* ( $p < 0.001$ ) significantly different from the contralateral leg for the entire sample

Compartment morphology results are presented in Table 3. A significant interaction (p < 0.001) revealed that although volume was smaller for both compartments on the ACLR compared to the contralateral leg, ST_dist_ was larger than ST_prox_ on the contralateral (p = 0.007), but not the ACLR (p = 0.369), leg. There were no differences in ACSA_max_ between compartments on either leg (main effect: p = 0.774; interaction: p = 0.951). However, a significant main effect (p = 0.002) revealed ACSA_max_ to be smaller in both ST compartments on the ACLR compared to the contralateral leg. As with volumetric results, a significant interaction (p = 0.002) revealed length to be shorter for both compartments on the ACLR compared to the contralateral leg, but ST_dist_ was longer than ST_prox_ on the contralateral (p < 0.001), but not the ACLR (p = 0.726), leg.

**Table 3.** Means and standard deviations of volume, maximal anatomical cross-sectional area (ACSA_max_), and length of proximal (ST_prox_) and distal (ST_dist_) semitendinosus compartments for contralateral and anterior cruciate ligament reconstructed (ACLR) legs. Repeated measures ANOVAs were performed for the entire sample and for the tendon regenerated only subgroup, and not for the non-regenerated only subgroup due to sample size. Only between-compartment statistics are presented below. Refer to the main text for main effects and interactions.

|  | Total (*n* = 10) |  |  |  | Regenerated only (*n* = 7) |  |  |  | Non-regenerated only (*n* = 3) |  |  |  |
| --- | --- | --- | --- | --- | --- | --- | --- | --- | --- | --- | --- | --- |
|  | Contralateral |  | ACLR |  | Contralateral |  | ACLR |  | Contralateral |  | ACLR |  |
|  | ST<sub>prox</sub> | ST<sub>dist</sub> | ST<sub>prox</sub> | ST<sub>dist</sub> | ST<sub>prox</sub> | ST<sub>dist</sub> | ST<sub>prox</sub> | ST<sub>dist</sub> | ST<sub>prox</sub> | ST<sub>dist</sub> | ST<sub>prox</sub> | ST<sub>dist</sub> |
| **Volume (cm<sup>3</sup>)** | 95.1 ± 25.1 | 116.3 ± 40.5** | 65.8 ± 26.2 | 71.7 ± 40.5 | 102.0 ± 27.2 | 131.2 ± 39.9## | 79.7 ± 16.5 | 91.5 ± 30.5 | 78.8 ± 7.4 | 81.4 ± 4.1 | 33.3 ± 0.7 | 25.3 ± 2.1 |
| **ACSA<sub>max</sub> (cm<sup>2</sup>)** | 9.4 ± 2.0 | 9.4 ± 2.3 | 7.1 ± 2.6 | 7.0 ± 2.9 | 10.0 ± 2.1 | 10.3 ± 2.2 | 8.4 ± 1.9 | 8.5 ± 2.2 | 8.1 ± 0.6 | 7.2 ± 0.2 | 4.1 ± 0.6 | 3.8 ± 0.4 |
| **Length (cm)** | 20.3 ± 2.4 | 23.7 ± 3.0*** | 18.5 ± 2.2 | 18.1 ± 5.8 | 20.5 ± 2.9 | 24.5 ± 3.3## | 19.5 ± 1.7 | 20.9 ± 4.3 | 19.8 ± 1.1 | 21.9 ± 0.4 | 16.1 ± 1.3 | 11.5 ± 1.9 |

\*\* (p < 0.01), \*\*\* (p < 0.001) significantly different from ST<sub>prox</sub> for the entire sample

The proximodistal length of the TI was shorter after ACLR (contralateral: 10.6 ± 2.0 cm; ACLR: 9.3 ± 2.0 cm; p = 0.005) but traversed a greater percent of muscle belly length (contralateral: 31.8 ± 5.7%; ACLR: 34.7 ± 4.7%; p = 0.015). The TI spanned from 28.8 ± 2.5% (TI_prox_) to 60.5 ± 4.7% (TI_dist_) of muscle belly length on the contralateral leg, and from 34.5 ± 7.2% (TI_prox_) to 69.2 ± 8.0% (TI_dist_) of muscle length on the ACLR leg. This corresponded to 47.6 ± 5.9% of ST_prox_ length to 44.3 ± 7.1% of ST_dist_ length on the contralateral leg and 49.4 ± 6.4% of ST_prox_ length to 53.2 ± 8.7% of ST_dist_ length on the ACLR leg. The location of ST_prox_ ACSA_max_ (contralateral: 29.6 ± 2.0%; ACLR: 35.7 ± 6.8%) did not differ from TI_prox_ on either leg (main effect: p = 0.126; interaction p = 0.615), although ST_prox_ ACSA_max_ and TI_prox_ were both more distal on the ACLR leg (main effect: p = 0.005) (Figure 2). While the location of ST_dist_ ACSA_max_ with respect to muscle length did not differ between legs (contralateral: 58.6 ± 3.9%; ACLR: 62.5 ± 9.7%; p = 0.139), a significant interaction (p = 0.021) revealed the location of ST_dist_ ACSA_max_ and TI_dist_ differed only on the ACLR leg (p = 0.025), and not the contralateral leg (p = 0.211), due to a slightly more distal position (relative to muscle length) of TI_dist_ after ACLR (p = 0.007).

**Figure 2.**
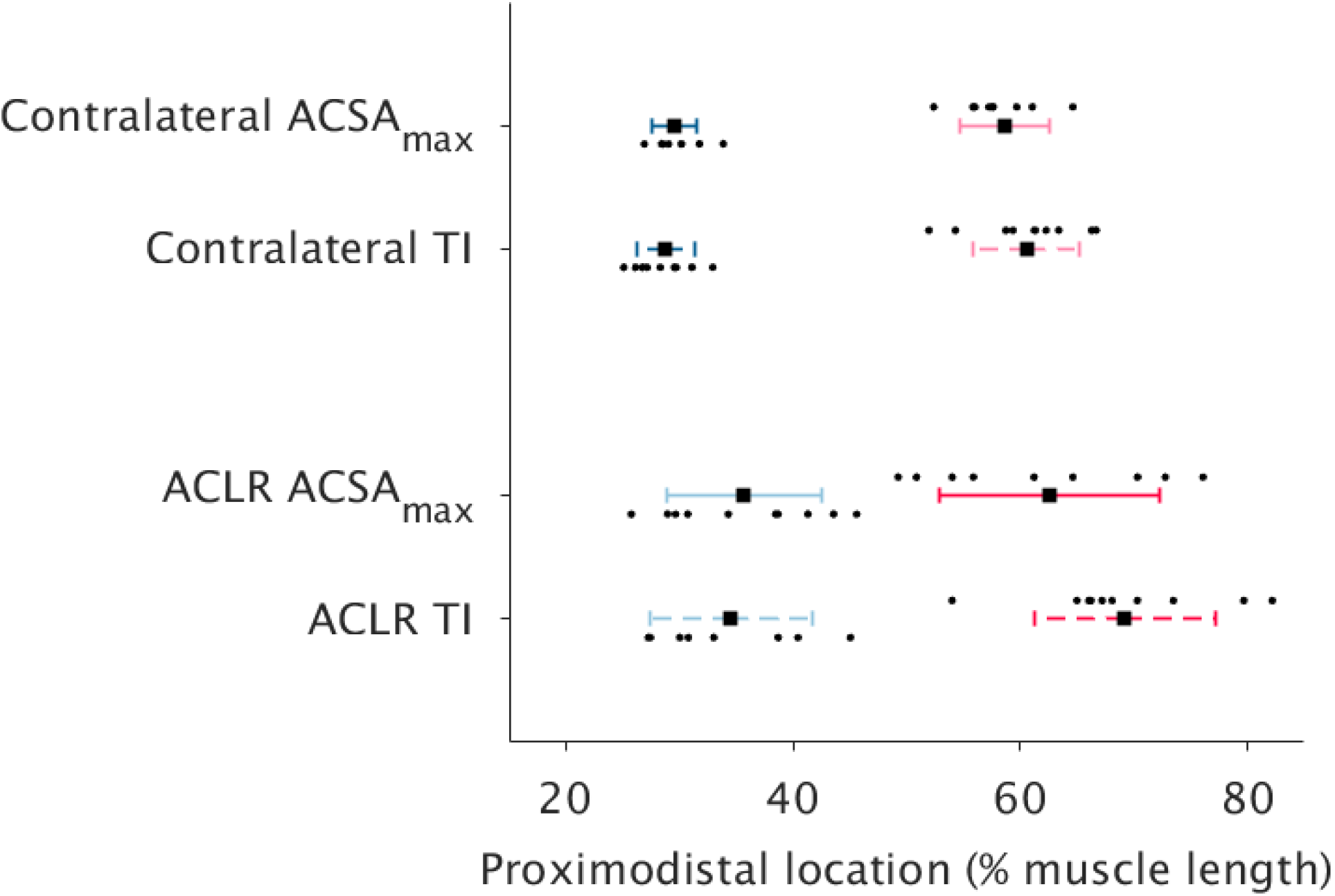
The location of proximal (contralateral: dark blue; anterior cruciate ligament reconstructed: light blue) and distal (contralateral: pink; anterior cruciate ligament reconstructed: red) semitendinosus compartment maximal anatomical cross-sectional area (ACSA_max_; unbroken lines) compared to tendinous inscription (TI) endpoints (broken lines) for anterior cruciate ligament reconstructed (ACLR) and contralateral legs. Data are presented as means and standard deviations, with dots representing individual data points.

The between-leg differences in each morphological parameter (volume, ACSA_max_, length) were highly correlated between compartments (*r* ≥ 0.66; p ≤ 0.037; Figure 3). Between-leg differences in compartment volume and ACSA_max_ were strongly correlated with corresponding whole ST muscle differences (*r* ≥ 0.93; p < 0.001; Figure 4), although length differences in ST_dist_ were much more strongly correlated with whole ST length differences (*r* = 0.99; p < 0.001) than was ST_prox_ (*r* = 0.75; p = 0.013). Conversely, the difference in ST_prox_ length was more strongly correlated (*r* = 0.99; p < 0.001) with the difference in TI length than was the length difference of ST_dist_ (*r* = 0.71; p = 0.021; Figure 5).

**Figure 3.**
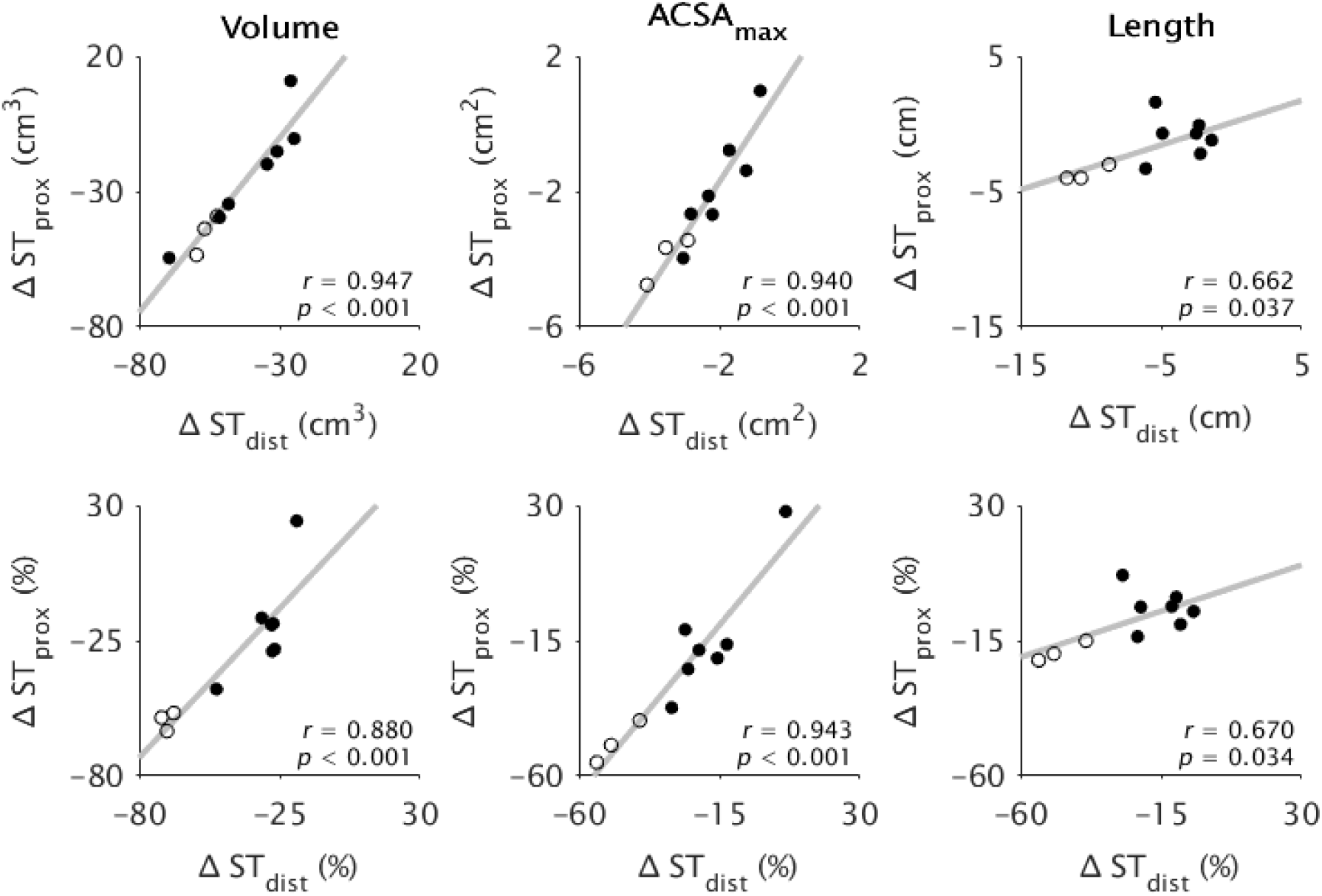
Pearson’s correlation coefficients (*r*) for the between-leg differences in proximal (ST_prox_) versus distal (ST_dist_) semitendinosus compartment volume (left), maximal anatomical cross-sectional area (ACSA_max_; middle), and length (right), plotted for absolute (upper) and relative (lower) differences. Dots represent individual data points from participants with (filled) and without (unfilled) tendon regeneration. All comparisons were significantly correlated (p ≤ 0.037).

**Figure 4.**
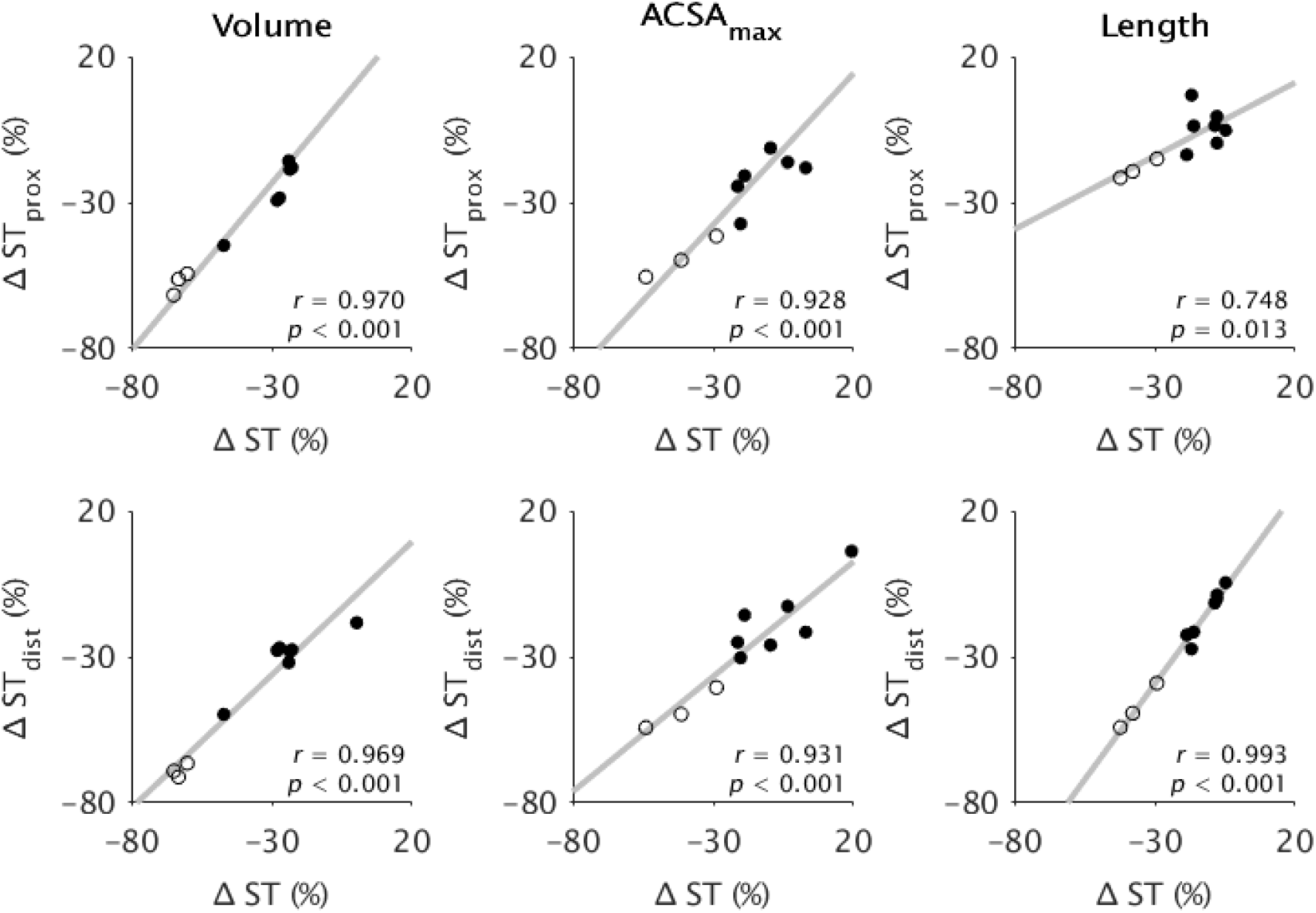
Pearson’s correlation coefficients (*r*) for the between-leg relative differences in whole semitendinosus (ST) muscle versus proximal (ST_prox_; upper) and distal (ST_dist_; lower) ST compartment volume (left), maximal anatomical cross-sectional area (ACSA_max_; middle), and length (right). Dots represent individual data points from participants with (filled) and without (unfilled) tendon regeneration. All comparisons were significantly correlated (p ≤ 0.013).

**Figure 5.**
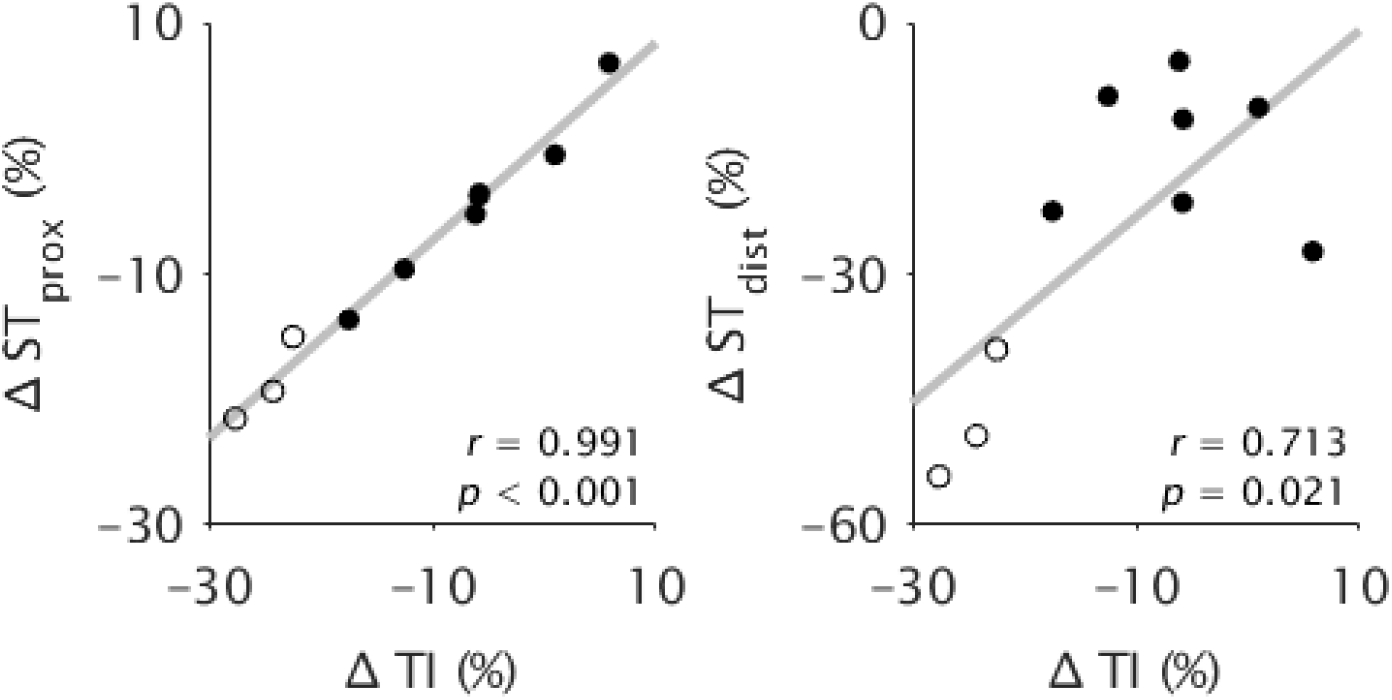
Pearson’s correlation coefficients (*r*) for the between-leg relative differences in tendinous inscription (TI) length versus between-leg relative differences in proximal (ST_prox_; left) and distal (ST_dist_; right) semitendinosus compartment length. Dots represent individual data points from participants with (filled) and without (unfilled) tendon regeneration. Both correlations were significant (p ≤ 0.021).

In the tendon regenerated subgroup, ST whole muscle volume (p = 0.007) and length (p = 0.002) differed between legs, although no statistical difference was detected for ACSA_max_ (p = 0.185). Compartment morphology results from the tendon regenerated subgroup demonstrated the same main effects and interactions as with the overall sample (Table 3). For volume, a significant interaction (p = 0.002) revealed ST_dist_ was larger than ST_prox_ only on the contralateral leg (contralateral: p = 0.004; ACLR: p = 0.185). For ACSA_max_, there was a main effect of leg (p = 0.031), but not compartment (p = 0.724), and no significant interaction (p = 0.551). For length, a significant interaction (p = 0.030) revealed ST_dist_ was longer than ST_prox_ only on the contralateral leg (contralateral: p = 0.002; ACLR: p = 0.261).

## DISCUSSION

This study is the first to characterize in detail the *in vivo* gross morphology of proximal and distal ST compartments in human adults, using ACLR patients as a model to study the morphological interactions between ST compartments. In the healthy contralateral leg, we found volume and length were both larger in ST_dist_ compared to ST_prox_, although ACSA_max_ did not differ between compartments. In the ACLR leg, no between-compartment differences in morphological parameters were seen, indicating larger volume and length changes in ST_dist_ compared to ST_prox_ following ACLR. We also found the TI endpoints to generally be positioned around the ACSA_max_ of each compartment. These results provide novel insight into the structure and function of the human ST muscle and how ST compartments contain the potential to be heterogeneously altered, particularly via their overall lengths.

### Gross morphology of semitendinosus

The ST compartment volumes from healthy contralateral legs were higher than previous reports due to the demographics studied previously (e.g., cadavers, children with or without diseased ST; Haberfehlner *et al*., [2016*b*, 2016*a*]; Hanssen *et al*., [2021]). We found the volume of ST_dist_ to be greater than ST_prox_ on the contralateral leg (Table 3). Previous studies (Haberfehlner *et al*., 2016*b*, 2016*a*; Hanssen *et al*., 2021) reported contradictory evidence to one another regarding any between-compartment volumetric differences, likely due to age and demographics. We also found ST_dist_ to be longer than ST_prox_, although it should be noted this refers to the proximodistal length of the respective compartment. Inferences regarding potential differences in fascicle length from our results are limited as the MRI sequences used only allow for gross morphology to be quantified. Nonetheless, a longer ST_dist_ compared to ST_prox_ is in agreement with studies of cats (Bodine *et al*., 1982; Edgerton *et al*., 1987; Loeb *et al*., 1987) and goats (Gans *et al*., 1989). The fiber and fascicle length of human ST_dist_ was originally reported to be longer than ST_prox_ (Markee *et al*., 1955; Barrett, 1962), although more recent dissections suggest average fascicle lengths may be equal between compartments (Wickiewicz *et al*., 1983; Woodley & Mercer, 2005; Kellis *et al*., 2012; Haberfehlner *et al*., 2016*b*). However, due to the oblique nature of the TI and muscle-tendon junctions, fascicle length can vary substantially proximodistally (Haberfehlner *et al*., 2016*b*) and depth-wise (Kellis *et al*., 2012) within a given compartment. In addition to three-dimensional freehand ultrasound, more complex MRI methods, such as diffusion tensor imaging (Bolsterlee *et al*., 2019), are needed to quantify compartment fascicle lengths *in vivo*.

Despite being unable to document at the level of fascicles, the between-compartment differences in length seem to explain the greater volume in ST_dist_ compared to ST_prox_, as ACSA_max_ did not differ between compartments. ACSA_max_, a strong determinant of muscle force, and by extension joint torque (Bamman *et al*., 2000; Fukunaga *et al*., 2001), is a good proxy of physiological cross-sectional area in muscles with little-to-no pennation, such as ST (Makihara *et al*., 2006; Haberfehlner *et al*., 2016*b*). Therefore, despite larger volume in ST_dist_, the lack of a between-compartment difference in ACSA_max_ suggests the maximal force producing capacity of each compartment does not differ in healthy legs. A lack of between-compartment differences in maximal force producing capacity (present study) and passive forces throughout the range of motion (Kositsky *et al*., 2022) support the paradigm of human ST_prox_ and ST_dist_ functioning mechanically as a single unit.

In accordance with previous reports (Lee *et al*., 1988; Woodley & Mercer, 2005; van der Made *et al*., 2015), we found the TI originated at approximately one-third of muscle length and continued obliquely into the lower half of the ST, although Garrett et al. (1989) found the TI to terminate slightly more proximally. We also found the TI endpoints (TI_prox_, TI_dist_) are centered approximately in the middle of each compartment, connecting regions where compartments are of their maximal size. Thus, the TI is well placed to interact between the two compartments, and the possible functional implications of this placement (e.g., force transmission) are discussed below (see *Role and function of the tendinous inscription*). Practically, TI endpoints coinciding spatially with compartment ACSA_max_ enables TI endpoints to be used as reference to standardize measures of maximal compartment size, which could also be performed using other, more accessible imaging modalities, such as ultrasonography (Haberfehlner *et al*., 2016*b*; Kositsky *et al*., 2020; Hanssen *et al*., 2021). However, given the slightly more proximal position of ST_dist_ ACSA_max_ compared to TI_dist_ in the ACLR leg, assessments of ACSA of ST_dist_ after ACLR should include images proximal to the end of the TI, to ensure ACSA_max_ is obtained. Additionally, as the location of whole ST muscle ACSA_max_ along the TI was highly variable, standardized locations for measures of ST ACSA_max_ (e.g., at 50% of TI length; Haberfehlner *et al*., [2016*b*]) should be taken with caution as potential inter-limb and/or inter-individual differences at that single location may just be normal variation.

### Effects of anterior cruciate ligament reconstruction on semitendinosus morphology

The differences in whole ST muscle morphology after ACLR were comparable with those seen in previous studies (Williams *et al*., 2004; Makihara *et al*., 2006; Snow *et al*., 2012; Nomura *et al*., 2015; Konrath *et al*., 2016; Messer *et al*., 2020). Although both compartments were smaller in volume and shorter after ACLR, the presence of statistically significant between-compartment differences in volume and length only for the contralateral, but not ACLR, leg indicates greater volume and length differences in ST_dist_ compared to ST_prox_ on the ACLR leg. Likewise, previous studies have also observed greater ST whole muscle volume loss in individuals with more muscle shortening (Williams *et al*., 2004; Nomura *et al*., 2015). Although de Moulin *et al*. (2022) found greater ACSA_max_ and volume differences in proximal and middle ST regions post-ACLR, regions were scaled based on whole muscle length. Thus, given the greater shortening in ST_dist_, it is highly likely that the results of de Moulin *et al*. (2022) were influenced by a different contribution of ST_prox_ and ST_dist_ to each muscle region between legs. A shorter and smaller ST_dist_ compared to ST_prox_ is consistent with findings in children with spasticity (Haberfehlner *et al*., 2016*a*). Greater differences in ST_dist_ compared to ST_prox_ post-ACLR is likely explained by the surgical procedure and the effects of epimuscular myofascial connections (Maas & Sandercock, 2010). To harvest the ST tendon, the sartorius fascia is incised and the most distal portion of the ST muscle belly is stripped off the ST tendon with a tendon harvester device. This surgical procedure not only damages the distal muscle end of ST_dist_, but further reduces the myofascial linkages that could maintain some loading through ST_dist_ and physically prevent muscle retraction in the absence of a distal insertion point. It is thus no surprise that attenuated strength deficit and less ST muscle shortening were found when only a partial width of the ST tendon was harvested (Sasahara *et al*., 2014), although future work is needed to determine if less severe morphological alterations in ST_dist_ would also have occurred with this partial ST tendon harvesting technique.

As fascicles of ST_prox_ terminate on the TI and a new set of fascicles (ST_dist_) originate on the TI (Markee *et al*., 1955; Barrett, 1962; Garrett *et al*., 1989; Woodley & Mercer, 2005; Haberfehlner *et al*., 2016*b*), the relationships between compartment shortening and whole muscle (Figure 4) and TI shortening (Figure 5) demonstrate the distoproximal manner of shortening consequent to distal tendon harvest (Street, 1983). The greater reduction in ST_dist_ compared to ST_prox_ length is likely due to initial retraction and the distal muscle stump being left free from distal attachment after tendon harvesting, and thus shortening of ST_dist_ is the immediate driver of whole ST length change. Shortening of ST_dist_ distoproximally would not independently have great influence on TI dimensions, as the TI is proximal to the main site of shortening. On the other hand, distoproximal shortening of ST_prox_, whose fascicles are distally attached to the TI, is thus the main regulator of TI shortening. Although we only measured its proximodistal length, the overall length of the TI would also have decreased due to the concomitant radial muscle atrophy. The shortening of the TI after ACLR may simply be slackening or crimping as a consequence of geometric constraints. On the other hand, as the mechanical behaviour of aponeuroses can change due to unloading (Lee *et al*., 2006) and aponeurosis width may increase in response to resistance training (Wakahara *et al*., 2015), TI shortening may also demonstrate the TI is a plastic structure that can adapt to its environment. The intrinsic modulation of the TI and whether these alterations occur concomitantly with, or delayed in response to, compartment changes should be examined in future investigations.

In contrast to different adaptations between compartments in volume and length and as was the case in the contralateral leg, ACSA_max_ did not differ between compartments in the ACLR leg, indicating the maximal force producing capacity was likely reduced by a comparable amount in both ST_prox_ and ST_dist_ after ACLR. The between-compartment relationships in the differences in morphological parameters (Figure 3) further demonstrate compartment morphology is linked, even after such drastic radial and longitudinal morphological adaptations. Large differences in ACSA_max_ between compartments would be severely detrimental to any mechanical interaction as transmission of high levels of force from a larger compartment to a smaller compartment would expose the latter to excessive stresses and a high risk of damage. The slightly different location of TI_dist_ compared to ST_dist_ ACSA_max_ post-ACLR may result from a change in the oblique angle of the TI, possibly leading to less efficient force transmission between compartments via altering the orientation between muscle fibers and the collagen within TI. Further, should greater proximodistal shortening of ST_dist_ be reflective of fascicle and sarcomere adaptations, the force-velocity characteristics of the two compartments may become incongruous. Additionally, the more drastic shortening of ST_dist_ may reflect a greater change in the length and/or number of sarcomeres in-series (Crawford, 1977; Abrams *et al*., 2000; Van Dyke *et al*., 2012), resulting in fibers in ST_dist_ being too short to produce high levels of force, particularly at highly flexed knee joint angles corresponding to short ST muscle belly lengths (Wickiewicz *et al*., 1984). A reduced operating range of ST is consistent with experimental results demonstrating greatly decreased knee joint moment in deep knee flexion post-ACLR with an ST graft (Makihara *et al*., 2006; Nomura *et al*., 2015; Morris *et al*., 2021). Future studies employing microendoscopy (Pincheira *et al*., 2022) may be valuable in elucidating compartment-specific changes at the level of the sarcomere across various joint angles in healthy and ACLR legs.

In the regenerated tendon subgroup, whole ST muscle ACSA_max_ did not statistically differ between legs. The failure to detect a significant difference in whole ST muscle ACSA_max_ in the regenerated tendon subgroup may stem from the large within-sample variation (between-leg mean difference -7.3 ± 14.9%), but may be a real finding given previous studies assessing ST ACSA_max_ post-ACLR did not statistically test this parameter for regenerated participants against a control or contralateral leg (Williams *et al*., 2004; Snow *et al*., 2012; Konrath *et al*., 2016; du Moulin *et al*., 2022) or used an average of five 3.6 mm slices when determining ACSA_max_ (Messer *et al*., 2020). Given ACSA_max_ of both ST compartments were significantly smaller on the ACLR leg in this subgroup, at minimum these results suggest measuring at the whole muscle level may not fully reflect adaptations at compartment level. Further, the longer and larger ST_dist_ compared to ST_prox_ in the contralateral leg only for both the whole sample and the regenerated tendon subgroup (Table 3) signifies having grouped all participants together did not influence the main results. Although we were unable to statistically compare between regeneration status due to the low sample size for this subgroup (n = 3), greater morphological differences seemed to occur in non-regenerated tendon individuals (Figures 3-5, Tables 2 and 3), which is consistent with previous reports of greater shortening and muscle atrophy after a lack of tendon regeneration (Davenport & Ranson, 1930; Crawford, 1977; Nomura *et al*., 2015; Konrath *et al*., 2016; du Moulin *et al*., 2022). Failure of the harvested distal tendon to regenerate would result in chronic underloading of ST_dist_, as seen by more extensive shortening of ST_dist_ in the non-regenerated cohort. Although the material and compositional properties of the regenerated tendon possibly differ from the native tendon (Papalia *et al*., 2015), the more substantial morphological changes when the ST tendon does not regenerate highlights the clinical and functional importance of facilitating tendon regrowth (when the whole tendon is harvested), as re-establishing a distal insertion point provides a functional mechanical linkage between the muscle, its surroundings, and bone, allowing greater loading and attenuation of ST atrophy. Future studies targeting improved ST tendon healing are thus welcome.

### Role and function of the tendinous inscription

The placement of an oblique, full-thickness TI within the human (and a variety of mammalian) ST has been puzzling anatomists for over a century (Humphry, 1869; Parsons, 1898). Ontogenetically, it is thought the TI marks the fusion between two separately developing anlagen (Macalister, 1868; Bardeen, 1906) and is possibly a neomorph (Appleton, 1928) resulting from the crossing of two muscles (Haines, 1934). However, Parsons (1898) noted a TI is not always present at the union of two muscle heads and thus there may be further morphogenetic explanations. Parsons’ sentiments were supported by later works finding a small number of fascicles bridge the TI and course from ST_prox_ to distal tendon insertion (Markee *et al*., 1955; Loeb *et al*., 1987; Woodley & Mercer, 2005), and that a TI separating compartments can also be present even when ST_prox_ is itself divided into two (dorsal and ventral) heads (Roy *et al*., 1984). As fascicles generally terminate (ST_prox_) or originate (ST_dist_) on the TI and to date intrafascicularly terminating muscle fibers within the human ST have not been observed (Barrett, 1962; Woodley & Mercer, 2005), the TI potentially serves to simply connect in-series muscle fibers (Humphry, 1872; Trotter *et al*., 1995). Connecting serial muscle fibers through a TI allows fibers to be of various lengths and experience varying levels of strain (Loeb *et al*., 1987), which could reduce the risk of fiber damage without severely affecting the function of ST given its wide operating range (Peters & Rick, 1977; Cutts, 1989), and allow for deep-to-superficial subunits within a given compartment (Bodine *et al*., 1982; Chanaud *et al*., 1991; Kellis *et al*., 2012). However, intrafascicularly terminating muscle fibers have been found in other human muscles with TIs (e.g., rectus abdominis; Cullen & Brödel, [1937]; Woodley *et al*. [2007]) and within ST compartments in other mammals whose ST contains a TI (Loeb *et al*., 1987; Gans *et al*., 1989). Therefore, even if human ST fibers do span entire fascicles, the TI seems to have another, main functional role than to just connect serial fibers.

Using shear-wave elastography, we recently indirectly demonstrated passive forces do not differ between human ST compartments, although it was unclear if forces were independently but equally developed or transmitted from one compartment to the other, resulting in equilibrium across the whole muscle (Kositsky *et al*., 2022). Here, we document the TI is advantageously positioned to possibly assist in force transmission by connecting the largest regions of each compartment (Figure 2), and this placement generally remains after the substantial gross morphological changes induced by harvesting the ST tendon for ACLR. As efficient force transmission to from muscle fibers to the connective tissue network occurs through shear at fiber ends (Purslow, 2020), the oblique arrangement of the TI provides a geometrical design facilitating shearing at the junction between fiber and connective tissue that would not be possible if the TI was completely transverse or coursing in the fascicle direction. The consequence of such an anatomical arrangement could allow for re-distribution and transmission of forces across fascicles of each compartment, as suggested by Kellis et al. (2012). Further, the TI endpoints being located around each compartment’s ACSA_max_ ensures the two compartments are mechanically linked and provides the TI a wide area over which to distribute forces. In support of a force transmission role of the TI, muscle fiber-TI connections have been reported to be comparable with myotendinous junctions (Hijikata & Ishikawa, 1997), and the TI of other muscles, such as in the cat neck, has been shown to house and/or be surrounded by Golgi tendon organs and muscle spindles (Richmond & Abrahams, 1975*a*, 1975*b*). Should the TI of ST also contain these sensory receptors, detection of local forces by Golgi tendon organs (Maas *et al*., 2022) and muscle spindles (Smilde *et al*., 2016) combined with the potential for asynchronous activation (English & Weeks, 1987; Hutchison *et al*., 1989) and unequal strains (Markee *et al*., 1955; Edgerton *et al*., 1987) between compartments provides a mechanism by which the central nervous system could use the TI to regulate compartmental force and stiffness to control intercompartmental coordination and enable efficient force transmission between compartments. Future studies combining complex computational models assessing force transmission (Sharafi & Blemker, 2011; Zhang & Gao, 2012) and muscle fiber interaction with internal aponeuroses (Knaus *et al*., 2022) may be able to confirm the main functional role(s) of the TI.

### Limitations

We only used the contralateral, non-surgical leg as the healthy baseline/control. However, unlike the quadriceps, hamstring morphology on the injured leg remains unchanged following anterior cruciate ligament injury alone (Lorentzon *et al*., 1989; Kariya *et al*., 1989; Williams *et al*., 2004; Konishi *et al*., 2012) and after ACLR the morphology of ST on the non-injured leg does not differ compared to pre-surgical (Williams *et al*., 2004) and control (Morris *et al*., 2021; du Moulin *et al*., 2022) groups. Additionally, the substantial between-leg differences in morphology we found compare well with previous literature (Williams *et al*., 2004; Makihara *et al*., 2006; Nomura *et al*., 2015; Konrath *et al*., 2016; Messer *et al*., 2020) and exceed bilateral asymmetry measures previously reported for ST (Williams *et al*., 2004; Kulas *et al*., 2018; Speedtsberg *et al*., 2022). Therefore, using the contralateral, non-injured leg as the baseline control was unlikely to have influenced the results. Further, compartment length was quantified by the proximodistal length of the respective compartment. As the TI is a complex three-dimensional structure, compartments are comprised of fascicles of various lengths (Kellis *et al*., 2012; Haberfehlner *et al*., 2016*b*) and thus compartment length may not accurately represent fiber or fascicle length. Therefore, we do not make any concrete conclusions at length scales below proximodistal compartment length as they were not possible to assess from our MRI scans. Finally, the ACLR surgical intervention induces secondary trauma at the knee joint and is thus more complex than regular tenotomy. However, slightly greater changes in ACSA are seen in the distal compared to proximal gracilis muscle when its distal tendon is harvested for shoulder reconstruction (Flies *et al*., 2020). Therefore, the results found in the present study are likely due to the ST tendon harvest for the ACLR procedure, rather than post-ACLR immobilization and disuse, but should be confirmed in future studies assessing ST compartment alterations after an ST tendon autograft has been used for reconstructing other lower (Cody *et al*., 2018; Stenroos & Brinck, 2020) and upper (Virtanen *et al*., 2014; Ranne *et al*., 2020) limb tendons. The compartment alterations in the ACLR leg may also not be representative of other (un)loading conditions, whose adaptations can also be assessed using the MRI acquisition parameters presented in this study.

## CONCLUSIONS

The proximal and distal compartments of human ST muscle appear to be modified in a non-uniform manner following harvest for ACLR. However, the heterogenous changes in length do not affect the homogeneity in compartment maximal radial size. The location of the tendinous inscription with respect to compartment morphology provides a wide area over which this connective tissue sheath could mediate the mechanical interaction of ST compartments. Overall, these results suggest the proximal and distal compartments of the human ST muscle are not mechanically independent.

## Data Availability

All data produced in the present study are available upon reasonable request to the authors

## Notes

### Competing Interest Statement

The authors have declared no competing interest.

### Funding Statement

This work was supported by a Griffith University Postgraduate Research Scholarship (to AK), the Erasmus+ progamme of the European Union (to AK and LS), and the Academy of Finland (#324529 to RKK, #332915 to LS).

### Author Declarations

The Human Research Ethics Committee of Griffith University gave ethical approval for this work.

